# Cardiac Troponin Fragmentation After Heavy Physical Exercise The MaraCat2 Study

**DOI:** 10.1101/2024.05.07.24307024

**Authors:** K. E. Juhani Airaksinen, Tuomas Paana, Tapio Hellman, Tuija Vasankari, Selma Salonen, Tuulia Tuominen, Konsta Teppo, Samuli Jaakkola, Olli J. Heinonen, Saara Wittfooth

## Abstract

**Background:** Elevations of commercial cardiac troponin T (total cTnT) levels are common after strenuous exercise, but there is limited information on whether the troponin composition differs from myocardial infarction (MI).

**Methods:** Troponin composition was analyzed in heparin plasma samples taken from 45 runners <1 h after marathon race and from 45 patients with Type 1 MI <12 h after the pain onset. The concentration of long molecular forms of cTnT (long cTnT) was measured with a novel upconversion luminescence assay, total cTnT was measured with a commercial high-sensitivity cTnT assay, and the ratio of long to total cTnT (troponin ratio) was determined.

**Results:** Total cTnT exceeded the upper reference limit (>14 ng/l) in 37 (82%) runners. The median total and long cTnT concentrations and the troponin ratio were higher (113 ng/l vs. 25 ng/l, 57 ng/l vs. 4.1 ng/l, and 0.36 vs. 0.17, respectively) in patients with MI than in marathon runners (p <0.001 for all comparisons). Troponin ratio decreased (r = -0.497, p <0.001) in marathon runners and increased (r = 0.466, p = 0.001) in patients with MI with increasing troponin release. In the receiver operating characteristics curve analyses of all subjects with cTnT release >14ng/l and those with 15-100 ng/l, long cTnT showed good predictive power with the area under the curve 0.969 (CI95% 0.939-1.000) and 0.937 (95% CI, 0.877–0.996) in discriminating marathon runners from MI patients. The respective values for total cTnT were 0.892 (CI95% 0.823-0.960) and 0.778 (95% CI, 0.656–0.901).

**Conclusions:** In contrast to Type 1 MI, after strenuous exercise only a small fraction of circulating cTnT exists as intact cTnT or long molecular fragments. The difference in troponin composition could be of diagnostic value when evaluating post-exercise cTnT elevations in subjects with chest complaints.

**Registration:** URL: https://www.clinicaltrials.gov; Unique identifier: NCT06000930

**Clinical Perspective:** *What Is New?:*

- Short cardiac troponin T (cTnT) fragments were the predominant form of cTnT release after marathon race among 45 runners in contrast to 45 patients with Type 1 myocardial infarction (MI) where intact cTnT and longer fragments are more commonly found in the circulation during the early hours after the attack also in patients with comparable troponin levels.
- The proportion of short cTnT components decreased (r = -0.497, p <0.001) in marathon runners and increased (r = 0.466, p = 0.001) in patients with MI with increasing troponin release.

*What Are the Clinical Implications?:*

- The principle of the novel immunoassay could be applied on automated platforms to allow implementation in clinical care and separate benign cTnT elevations after strenuous exercise from those of acute MI in a single sample with better accuracy than the commercial high-sensitivity cTnT test.

---

Cardiac troponins are in a key role in the diagnosis of acute myocardial infarction (MI).^1^ Minor cardiac troponin elevations are common also after strenuous exercise, causing diagnostic challenges when associated with chest discomfort.^2,3^ The exact mechanisms of cardiac troponin rise after strenuous physical activity remains ill-defined, but according to a small gel filtration chromatography study, the released cardiac troponin T (cTnT) in this condition seems to be predominantly in the form of small molecules.^4^ Temporary increase in permeability of cell membranes may allow the leakage of these smaller cytosolic troponin fragments to circulation.

Importantly, the current commercial high-sensitivity cTnT test detects the small and long troponin fragments and the intact cTnT containing the stable central part of the cTnT molecule and is called here the total cTnT assay.

In a Proof-of-Principle study, we developed a simple time-resolved immunofluorometric assay based on europium chelate labels for the measurement of long cTnT molecules. The assay showed a high accuracy in discriminating between cTnT elevations in MI and end-stage renal failure.^5^ In the present study protocol, we compared the characteristics of troponin release after a marathon race and Type 1 MI using an improved version of the long cTnT test and the commercial total cTnT test.

## Methods

### Patients and samples

A total of 45 recreational runners (31 male) aged 28-75 (median 35) years participating in the Paavo Nurmi Marathon 2023 in Turku were recruited to the MaraCat2 Study (ClinicalTrials.gov Identifier:

NCT06000930) with an open email invitation. Eighteen participants finished the full marathon and 27 the half-marathon. None of the patients had a history of coronary artery disease. All participants gave a lithium-heparin plasma sample within 60 min after finishing the race.

A control group of 45 patients (36 male) with acute MI (25 ST elevation MI) were recruited among patients admitted to Heart Centre of Turku University Hospital (ClinicalTrials.gov Identifiers: NCT04465591 and NCT05858112). Coronary angiography was performed in all included patients to confirm culprit lesion and the MI diagnosis and all included patients were treated with primary or urgent percutaneous coronary intervention. Only patients with a delay less than 12 hours from symptom onset to lithium-heparin plasma sample collection and estimated glomerular filtration rate > 30 mL/min/1.73m^2^ were included.

All participants provided a written informed consent. The study complies with the Declaration of Helsinki as revised in 2013 and the study protocol was approved by the Medical Ethics Committee of the Hospital District of Southwest Finland.

The samples were analyzed fresh for total cTnT. In separate sample tubes the plasma was aliquoted after centrifugation and stored at −70 °C until analysis by the long cTnT assay.

### Analytical methods

The novel highly sensitive two-step heterogenous sandwich-type immunoassay using upconversion luminescence for signal production was used for the detection of long molecular forms of cTnT.^6^ The anti-cTnT monoclonal antibodies (mAb) and human cardiac troponin ITC-complex used as a calibrator were obtained from HyTest Ltd (Turku, Finland). The capture antibody (7E7 mAb) and the tracer antibody (1C11 mAb) bind to amino acid residues (aar) 223–242 and 174–190 of cTnT, respectively. The C-terminal region of cTnT between these two epitopes (aar 190–223) contains several cleavage sites and thus, the ability of the assay to detect long forms of cTnT is based on targeting the cTnT molecules that are not degraded at aar 190–223. The limit of detection and limit of quantitation of this assay are 0.4 ng/l and 1.8 ng/l, respectively^6^.

Total cTnT was analyzed using a commercial high-sensitivity cTnT assay (Elecsys hs-cTnT, Roche Diagnostics GmbH, Mannheim, Germany). For this assay, the limits of detection and quantitation are 3 ng/l and 13 ng/l, respectively. The ratio of long cTnT forms/total cTnT (troponin ratio) was used as the measure of troponin fragmentation.^5^

### Statistical analysis

Continuous variables are reported as median (25^th^ – 75^th^ percentiles) and categorical variables as counts (percentage). Mann–Whitney *U* test was used for group comparisons. Chi-squared test and Fisher’s exact test were used for categorical variables as appropriate. Correlation between continuous variables was estimated using the Spearman’s test. Linear regression analysis was used to identify factors significantly relating to total and long cTnT levels and their ratio in marathon runners and MI patients. All predictors with a p value < 0.1 in univariate analysis were included in the final regression model. Receiver operating characteristics (ROC) curve analyses were performed to estimate the area under the curve (AUC) to measure the discriminative capacity of cTnT, long cTnT and the troponin ratio between marathon runners and MI patients. Additional analyses were performed to assess their discriminative capacity between marathon runners and MI patients with cTnT levels ranging from 15 to 100 ng/l, the cTnT level considered to be most important in the differential diagnosis between MI and exercise-induced cTnT release. All tests were two-sided, and the limit of significance was set at p < 0.05. Sigmaplot 15 (Inpixon, California) was used for the comparison of ROC curves and IBM SPSS Statistics software version 26.0 was used to perform all other analyses.

## Results

### Total cTnT, long cTnT, and troponin ratio in marathon runners and patients with myocardial infarction

None of the runners reported cardiac symptoms after the race. Total cTnT exceeded the upper reference limit (>14 ng/l) in 37 (82%) runners with a median concentration of 25.0 ng/l (range 7-63 ng/l). Median long cTnT concentration was 4.1 ng/l (range 1-8.4 ng/l) (Table 1). The total and long cTnT levels were higher after full marathon than after half-marathon (33 (25-47 ng/l) vs. 20 (11-27 ng/l) and 5.6 (3.6-7.6 ng/l) vs. 3.3 (2.2-4.7 ng/l), p = 0.002 for both comparisons). Troponin ratio decreased significantly (r = -0.497, p <0.001) with increasing total cTnT release (Figure 1).

**Table 1.**
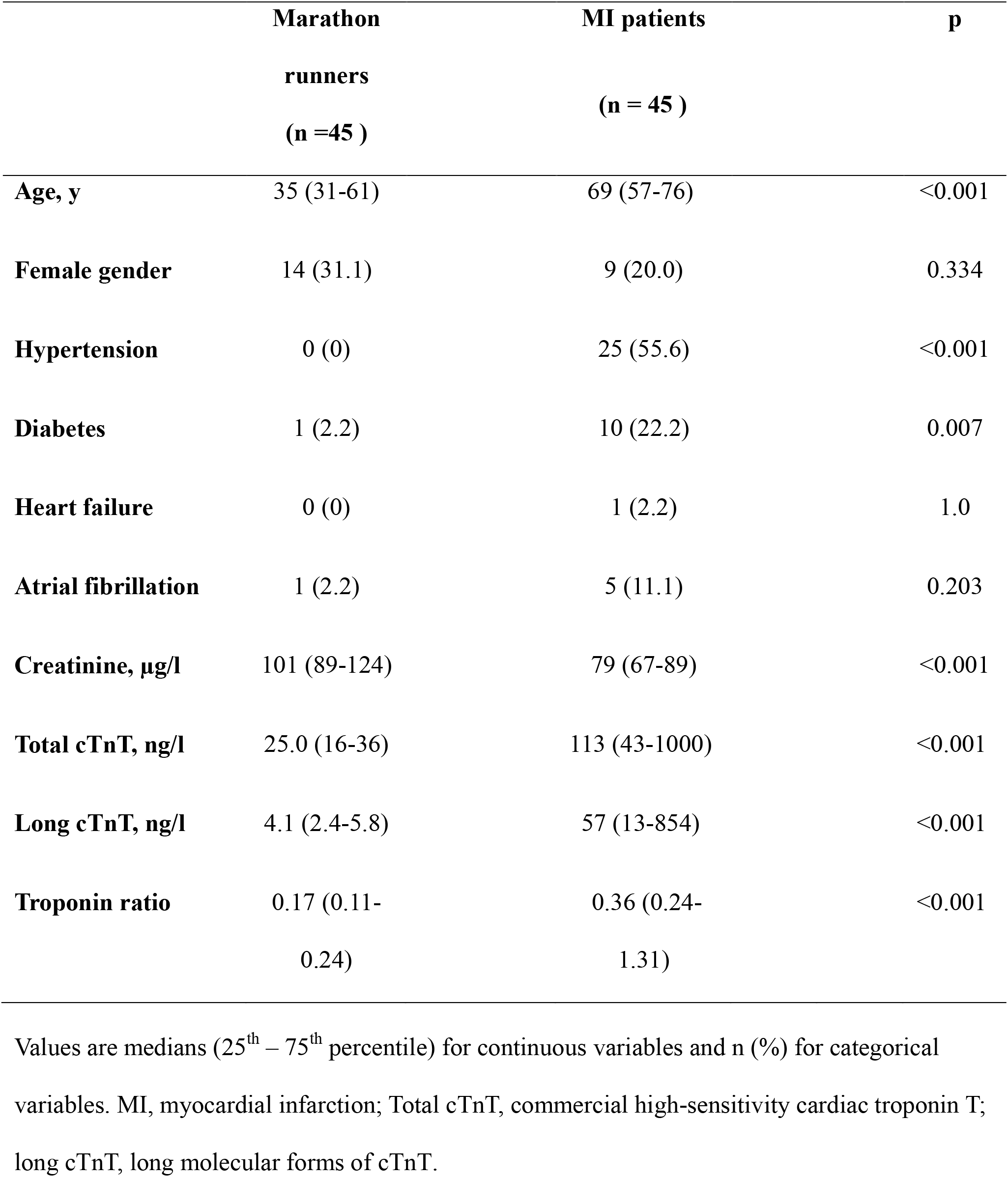
Clinical Characteristics and Total and Long cTnT and Their Ratio (Troponin Ratio) in the Study Groups.

|  | **Marathon**<br>**runners**<br>**(n=45 )** | **MI patients**<br>**(n = 45 )** | **p** |
| --- | --- | --- | --- |
| **Age, y** | 35 (31-61) | 69 (57-76) | <0.001 |
| **Female gender** | 14 (31.1) | 9 (20.0) | 0.334 |
| **Hypertension** | 0 (0) | 25 (55.6) | <0.001 |
| **Diabetes** | 1 (2.2) | 10 (22.2) | 0.007 |
| **Heart failure** | 0 (0) | 1 (2.2) | 1.0 |
| **Atrial fibrillation** | 1 (2.2) | 5 (11.1) | 0.203 |
| **Creatinine, µg/l** | 101 (89-124) | 79 (67-89) | <0.001 |
| **Total cTnT, ng/l** | 25.0 (16-36) | 113 (43-1000) | <0.001 |
| **Long cTnT, ng/l** | 4.1 (2.4-5.8) | 57 (13-854) | <0.001 |
| **Troponin ratio** | 0.17 (0.11-0.24) | 0.36 (0.24-1.31) | <0.001 |

Values are medians (25<sup>th</sup> – 75<sup>th</sup> percentile) for continuous variables and n (%) for categorical variables. MI, myocardial infarction; Total cTnT, commercial high-sensitivity cardiac troponin T; long cTnT, long molecular forms of cTnT.

**Figure 1.**
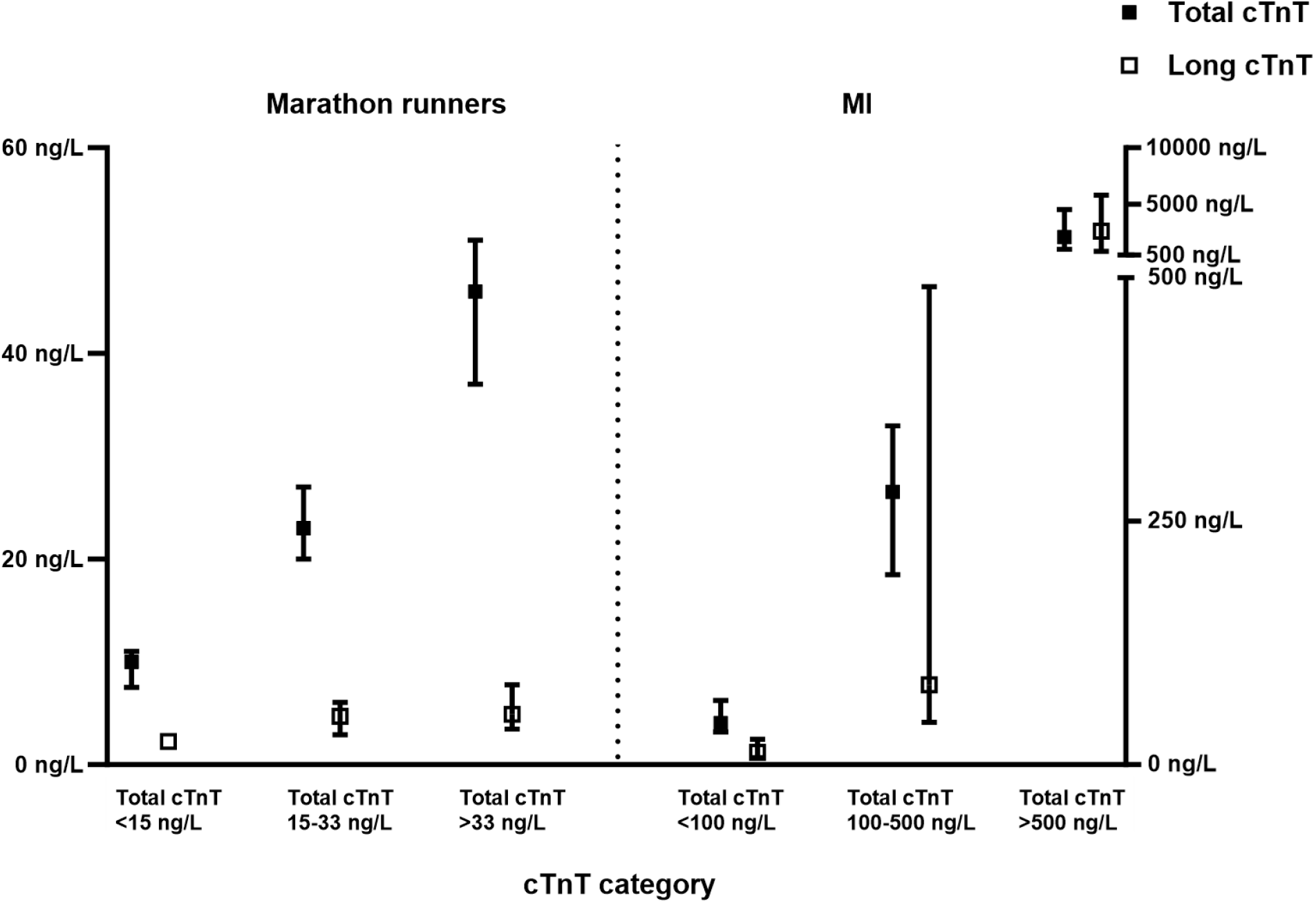
Concentrations of long forms of cTnT (long cTnT) in relation to the magnitude of total cTnT release. The share of long cTnT of total cTnT (troponin ratio) decreased (r = -0.497, p <0.001) with increasing total cTnT release in marathon runners and increased (r = 0.466, p =0.001) in patients with myocardial infarction (MI).

Both the total cTnT and long cTnT concentrations as well troponin ratio were higher (p<0.001 for all) in MI patients than in marathon runners (Table 1). In contrast to the marathon runners, in MI patients troponin ratio increased significantly (r = 0.466, p = 0.001) with increasing total cTnT release (Figure 1). Total cTnT and long cTnT concentrations and troponin ratio were not related to the time delay from pain onset to blood sampling or any clinical features of the MI patients listed in Table 1.

### Diagnostic performance of long cTnT measurement

To assess the diagnostic value of long cTnT, we compared the troponin composition of the 37 marathon runners having elevated total cTnT levels (>14 ng/l) with all MI patients and with a subgroup of MI patients (N=22) having total cTnT less than 100 ng/l. In both of these analyses, total cTnT, long cTnT and troponin ratio were higher (p<0.001) in patients with MI (Figure 2). In the ROC curve analyses, both the total and long cTnT showed excellent predictive power in discriminating marathon runners from MI patients with a small but statistically significance difference in favor of long cTnT, with respective AUCs of 0.892 (CI95% 0.823-0.960) and 0.969 (CI95% 0.939-1.000) (p = 0.012). The cutoff point of 8.2 ng/l for the long cTnT ratio showed a sensitivity of 86.7% ja specificity of 97.3% in separating runners and MI patients (Figure S1).

**Figure 2.**
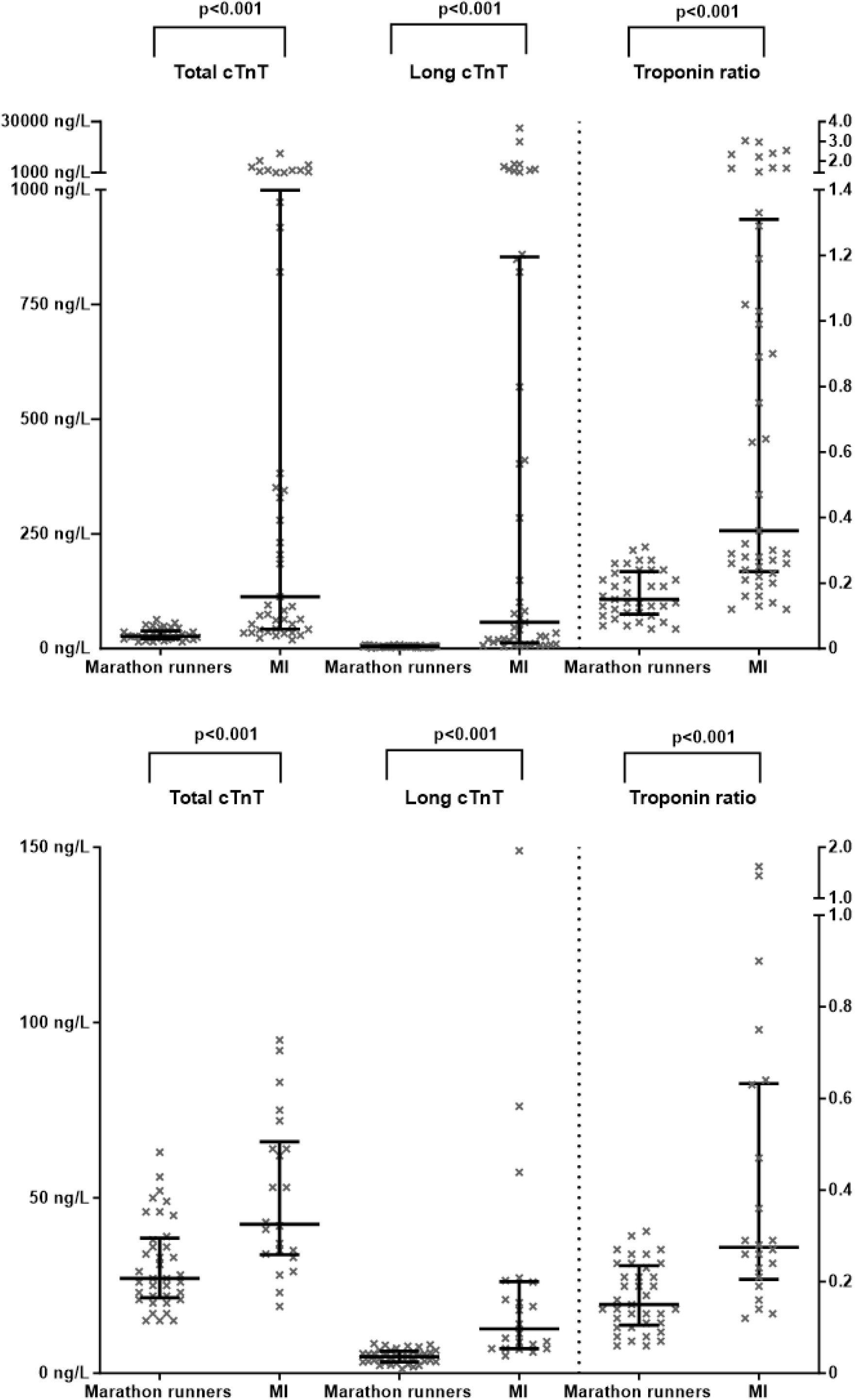
Immunoassay for long cTnT forms to discriminate between marathon runners with total cTnT >14ng/L and patients with MI. Total cTnT, long cTnT forms (Long cTnT), and troponin ratio (ratio of long cTnT forms to total cTnT) in marathon runners with total cTnT >14ng/l and in patients with myocardial infarction (MI) and <12-hour delay between symptom onset and blood sampling (Upper Panel) and in a subgroup of patients with total cTnT 15-100 ng/l (Lower Panel).

When restricting the analysis only in subjects with total cTnT concentration from 15 to 100 ng/l, the respective AUC values for total cTnT and long cTnT were 0.778 (95% CI, 0.656–0.901) and 0.937 (95% CI, 0.877–0.996) (p = 0.007).

## Discussion

As expected, the majority of runners had post-race cTnT levels above the rule-in criteria for the diagnosis of acute MI and the troponin release was more marked after running the full marathon. Most importantly, we showed that short cTnT fragments are the predominant form of cTnT release after marathon race in contrast to Type 1 MI where longer and intact cTnT forms are more commonly found in the circulation during the early hours after MI also in patients with comparable troponin levels.

The present findings on troponin fragmentation after exercise are in line with the gel filtration chromatography study by Vroeman et al showing in 10 runners with a postrace cTnT exceeding 70 ng/l that this cTnT release concerned only secondary cTnT fragments of which the N-terminal and C-terminal ends of the protein were cleaved off.^4^ The results with our sensitive immunoassay technique confirmed this in a larger group of runners and with less profuse troponin release.

Importantly, we could show that the magnitude of long cTnT release remained at a low level in spite of increasing release of total cTnT. The current finding on smaller troponin fragments as the predominant component of circulating cTnT after exercise is similar to the troponin composition seen in patients with end-stage renal failure, although the long cTnT levels in renal patients were slightly lower than the present concentrations after exercise.^5,7^ In MI, intact and longer forms of troponin were the predominant form of troponin release in patients with extensive myocardial damage, which is in line with previous reports on small groups of patients with large ST elevation MI.^5,8,9^ As a novel finding, our sensitive test showed that the troponin composition was related to the magnitude of myocardial injury and the cTnT release included more smaller components in patients with milder myocardial injury.

Proposed mechanisms for exercise-induced cTnT elevations include increased cell membrane permeability or cellular release of proteolytic troponin degradation products during exercise.^3^

Enhanced myocyte turnover, necrosis, and apoptosis are among other potential mechanisms of cardiac troponin release. The role of temporary increase in cell membrane permeability is supported by small cardiac magnetic resonance imaging studies which have detected myocardial oedema, transient increases in mean diffusivity and extracellular volume with a positive correlation to troponin release after marathon race.^10,11,12^

In clinical practice problems may arise when subjects present with chest pain to the emergency department shortly after strenuous exercise and have elevated troponin levels. Electrocardiogram is often normal, but in some cases athlete’s heart may complicate the interpretation. The present observations on marked differences in troponin composition between Type 1 MI and exercise-induced benign troponin release could be helpful in the diagnostic workup of the patients in the “grey zone” of mildly elevated troponin release.

Earlier research on troponin fragmentation has employed gel filtration chromatography, Western blotting and mass spectrometry, techniques that are too complicated for clinical use.^4,8,9^ The low analytical sensitivity of these methods is another obstacle for clinical applications.^4,9^ In contrast, our immunoassay approach is a significantly more sensitive method for the analysis of cTnT fragmentation. Importantly, the principle of our assay could be applied on automated platforms to allow implementation in clinical care to improve the accuracy and rapidity of laboratory diagnostics of MI.

Our study group was quite small and the troponin release after the race was less than in many previous reports. We did not collect blood samples before the race, but based on earlier research, chronic elevation of troponins in these healthy subjects is unlikely.^2,3^ In some patients the long cTnT concentrations were higher than the total cTnT values in our study. This is likely due to differences in the calibration of the assays. It should be noted, that fragmentation of troponins seems to be a continuous process after MI.^5,13^ Longer delays between MI symptom onset and sampling are likely to dilute the observed differences in troponin composition.

In conclusion, this novel highly sensitive long cTnT immunoassay shows that the troponin release after strenuous exercise is composed mainly of smaller troponin fragments. The test holds promise that measuring long cTnT forms could help to separate benign cTnT elevations after strenuous exercise from those of acute MI in a single sample with better accuracy than the commercial high-sensitivity cTnT test.

## Data Availability

Details of the patients, methods, and data supporting the present findings are available from the corresponding author on reasonable request.

## Acknowledgements

We would like to thank Jaana Rosenberg from the University of Turku for providing us with the upconverting nanoparticle labels.

## Sources of Funding

This work was supported by the Finnish Foundation for Cardiovascular Research, Helsinki, Finland, Clinical Research Fund (EVO) of Turku University Hospital, Turku, Finland, the Finnish Society of Clinical Chemistry, Helsinki, Finland, the Turku University Foundation, Turku, Finland, and the Varsinais-Suomi Regional Fund of the Finnish Cultural Foundation, Turku, Finland.

The funding organizations had no role in the design and conduct of the study; collection, management, analysis, and interpretation of the data; preparation, review, or approval of the manuscript; and decision to submit the manuscript for publication.

## Disclosures

K.E. Juhani Airaksinen: Research grants from the Finnish Foundation for Cardiovascular Research and Clinical Research Fund of Turku University Hospital, Turku, Finland. Lectures for Astra Zeneca, Bayer, Boehringer Ingelheim, Pending patent application WO2023187258 (A1) - ASSAY FOR LONG FORMS OF CARDIAC TROPONIN T

Tuomas Paana: Lectures for Astra Zeneca

Tuija Vasankari: Pending patent application WO2023187258 (A1) - ASSAY FOR LONG FORMS OF CARDIAC TROPONIN T

Selma Salonen: None

Tuulia Tuominen: None

Tapio Hellman: Lectures for AstraZeneca, Astellas and GSK, Pending patent application WO2023187258 (A1) - ASSAY FOR LONG FORMS OF CARDIAC TROPONIN T

Konsta Teppo: Research grants from The Finnish Foundation for Cardiovascular Research, Aarne and Aili Turunen Foundation, The Finnish Medical Foundation, The Finnish Foundation for Alcohol Studies and the Finnish State Research Funding.

Samuli Jaakkola: Lectures for Amgen, Boehringer Ingelheim, BMS Pfizer

Olli J. Heinonen: None

Saara Wittfooth: Research grants from the Finnish Society of Clinical Chemistry, the Turku University Foundation and the Varsinais-Suomi Regional Fund of the Finnish Cultural Foundation, and the Finnish Foundation for Cardiovascular Research. Research funding from Business Finland, official Finnish government agency for trade and investment promotion, innovation funding, travel promotion and talent attraction. Pending patent application WO2023187258 (A1) - ASSAY FOR LONG FORMS OF CARDIAC TROPONIN T

## Non-standard Abbreviations and Acronyms

MI: Myocardial infarction
cTnT: Cardiac troponin T
mAb: monoclonal antibodies
aar: amino acid residues
AUC: Area under the curve
ROC: Receiver operating characteristics

---

**Figure S1.**
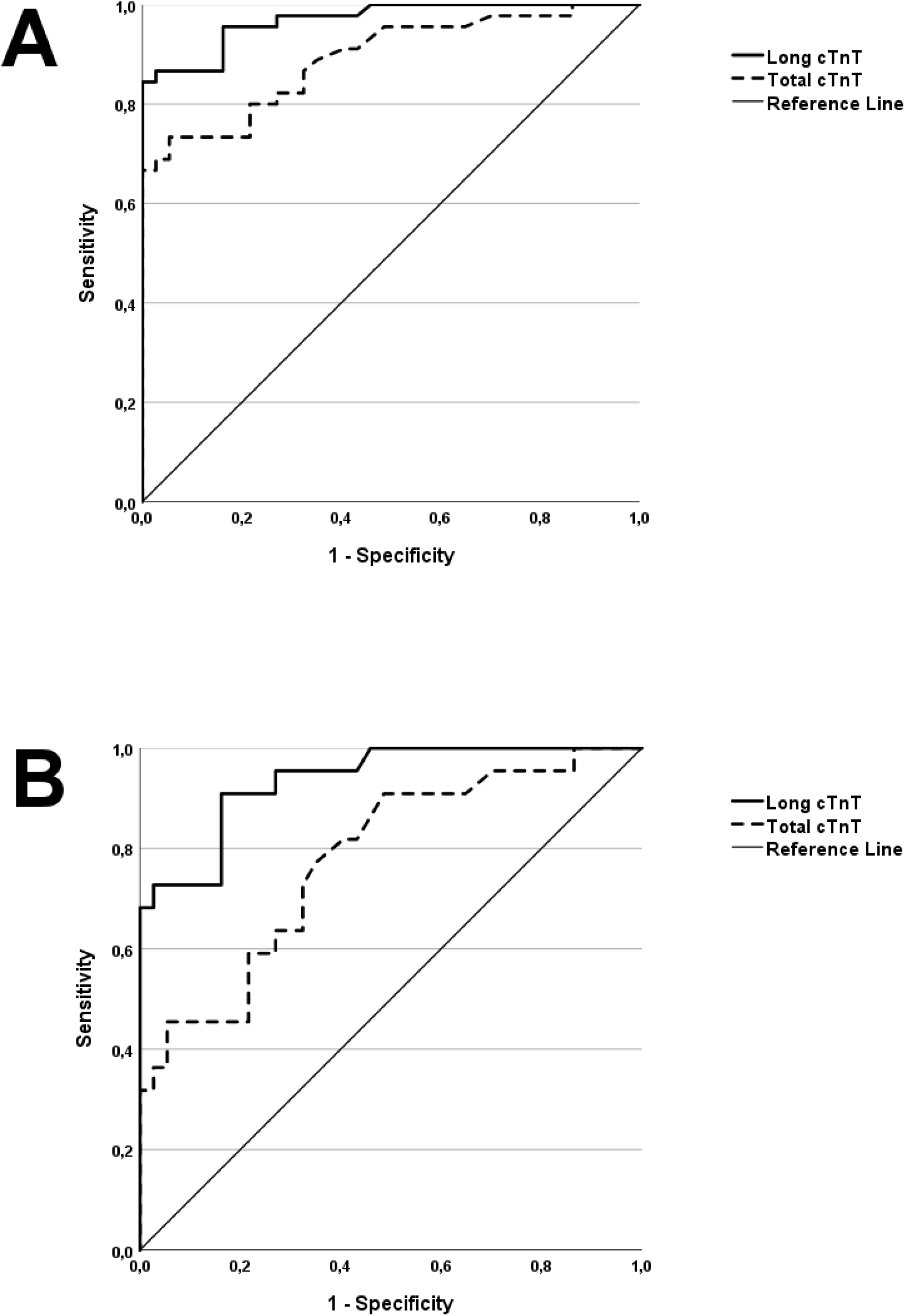
Receiver operating characteristic curves depicting the discriminative capacity of long cTnT (full line) and total cTnT (dotted line) between marathon runners with cTnT release >14ng/l and MI patients. (all patients: AUCs of 0.892 [CI95% 0.823-0.960] and 0.969 [CI95% 0.939-1.000] [panel A]; and MI patients with total cTnT release 15-100 ng/L: 0.778 [95% CI, 0.656– 0.901] and 0.937 [95% CI, 0.877–0.996] [panel B]). AUC indicates area under the curve; MI, acute Type 1 myocardial infarction with time from pain onset to blood sample < 12 hours; cTnT, cardiac troponin T.

## Notes

### Competing Interest Statement

The authors have declared no competing interest.

### Clinical Trial

https://www.clinicaltrials.gov; Unique identifier: NCT06000930

